# Assessment of Infection Prevention and Control Protocols, Procedures, and Implementation in Response to the COVID-19 Pandemic in Twenty-three Long-term Care Facilities in Fulton County, Georgia

**DOI:** 10.1101/2020.08.13.20174466

**Authors:** Carson T. Telford, Cyndra Bystrom, Teresa Fox, Sherry Wiggins-Benn, Meshell McCloud, David P. Holland, Sarita Shah

## Abstract

Through infection prevention and control (IPC) site visits to 23 LTCFs in Fulton County, Georgia, comparison between the Higher- and Lower-prevalence groups revealed significant differences in PPE and Social Distancing, with five specific indicators driving these differences.

## Background

Long-term care facility (LTCF) residents are among the populations at greatest risk of experiencing severe outcomes from Coronavirus Disease 2019 (COVID-19) infection (*1-2*). Fulton County, which covers the city of Atlanta, Georgia and is home to more than one million inhabitants, and as of July 29, 2020 has received reports of 1,188 COVID-19 infections in residents from 45 LTCFs within its jurisdiction. Among infected residents, approximately 22% were hospitalized and 15% died; 51% of COVID-19 deaths in Fulton County are attributed to LTCF residents (*3*).

At the beginning of the COVID-19 pandemic, the Centers for Disease Control and Prevention (CDC) developed interim infection prevention and control (IPC) recommendations for healthcare personnel for preventing COVID-19, which included specific guidelines for LTCFs (*4-6*). In response to outbreaks in LTCFs, the Fulton County Board of Health (FCBOH) organized a COVID-19 LTCF Outbreak Response Team in collaboration with the Georgia Department of Public Health to evaluate IPC strategies through site visits to LTCFs. The purpose was to provide support to Fulton County LTCFs by identifying gaps in IPC protocols and provide real-time feedback on how to effectively prevent additional infections, hospitalizations, and deaths due to COVID-19. We report the overall strengths and weaknesses in IPC protocols in participating LTCFs and provide recommendations for improving IPC practices.

## Methods

In June 2020, site visits to LTCFs in Fulton County were conducted by professionals in infection prevention, epidemiology, and nursing. Site selection for the visits prioritized LTCFs with the highest prevalence of COVID-19 infection; facilities with lower prevalence were also visited upon request. We defined LTCF as nursing homes, skilled nursing facilities, and assisted living facilities. Site visits were conducted in person or virtually by video conferencing, and, for some facilities, both. We limited the number of in-person consultants per visit to five or fewer, if requested by the LTCF, to allow for social distancing and to reduce the risk of introducing infection in a LTCK. Team members used personal protective equipment (PPE) and were screened for symptoms and elevated temperature in accordance with CDC recommendations, prior to entrance (*4*).

Thirty-three key indicators were evaluated from five IPC categories: Hand Hygiene, Disinfection, Social Distancing, PPE, and Screening for symptoms and elevated temperature; indicators were sourced from literature providing COVID-19 recommendations for prevention and control of COVID-19 in LTCFs (*4-7*). Frequency distributions (counts and percentages) were used to describe the overall LTCF adherence to each key indicator. All residents of LTCFs were tested in accordance with federal and state orders; the overall infection prevalence was calculated as the total number of infected residents divided by the total number of residents in facilities visited. A comparison analysis stratified LTCFs into two groups: those whose resident infection prevalence was higher than the overall prevalence of sites visited (Higher-prevalence group) and those with infection prevalence lower than the overall prevalence (Lower-prevalence group). Overall implementation of key indicators within each category was also calculated as a composite proportion.

Chi-square test of proportions was used to test differences between the Higher- and Lower-prevalence groups and p<0.05 was used as the cut-off for statistical significance. Statistical differences between the groups for continuous variables were calculated using a two-tailed T-test for two independent means, using p<.05 as the cut-off for statistical significance. Key indicators that were continuous variables (n=2) were not included in the composite implementation calculation for IPC categories.

This activity was reviewed by the Georgia Department of Public Health Institutional Review Board and deemed exempt from IRB review as a public health surveillance activity in response to the COVID-19 emergency response.

## Results

Data was gathered from site visits to 23 out of the 45 Fulton County LTCFs which reported ≥1 COVID-19 infection. These facilities accounted for 76%, 84%, and 83% of all reported cases, hospitalizations, and deaths in LTCF residents in Fulton County, respectively. The prevalence of resident infection in participating LTCFs was 37%, while the overall prevalence of all resident infections in Fulton County LTCFs with ≥ 1 infection in Fulton County was 29% (**Table 1**). The Higher-prevalence group’s resident infection proportion was 62% (Range: 46-74%), while the Lower-prevalence group’s resident infection proportion was 15% (Range: 1-33%; **Table 1**). The proportion of residents who were hospitalized and died among those who were infected was similar between both groups (**Table 1**).

**TABLE 1.**
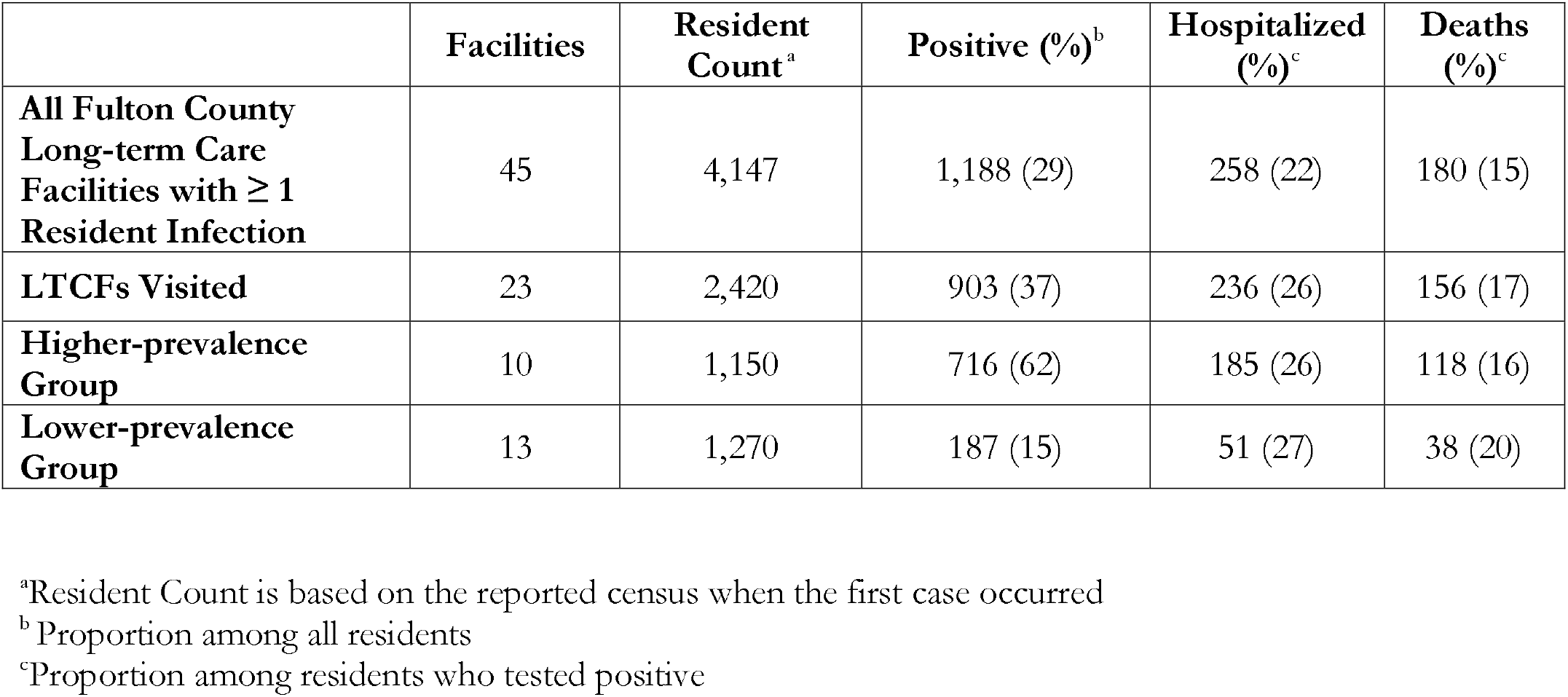
Prevalence of infection, hospitalization, and death due to COVID-19 in Long Term Care Facilities in Fulton County, Georgia.

Overall, IPC implementation was lowest in the Disinfection category (33%) and highest in the Screening category (75%). In the Disinfection category, 61% of LTCFs had a certified infection preventionist on staff and 26% were training and auditing staff on the proper use of cleaning products, including wet times and implementation of the two-step method. Cleaning logs documenting disinfection of shared items (i.e. IV polls, wheel chairs, shared blood pressure cuffs) were only present in 13% of LTCFs.

In the Hand Hygiene category, only 39% of LTCFs had hand sanitizer available in all essential locations (i.e., nursing stations, medical carts, outside COVID unit, in patient rooms). Protocols to enforce social distancing in small enclosed spaces such as elevators or PPE donning and doffing rooms were established in 35% of LTCFs. No LTCFs were implementing the Buddy System for donning and doffing PPE at the time of our site visit, which necessitates nursing staff observing each other through the PPE donning and doffing process.

Ten LTCFs had infection prevalence in residents greater than the overall infection proportion (37%) and were therefore classified as the “Higher-prevalence group” while the “Lower-prevalence group” comprised 13 LTCFs whose infection prevalence was lower than the overall proportion. Significant differences in implementation of IPC indicators between the Higher- and Lower-prevalence facilities were observed in the Social Distancing category (Higher-prevalence group 54% vs. Lower-prevalence group 72%, p=0.01; **Table 2**) and the PPE category (Higher-prevalence group 41% vs. Lower-prevalence group 72%, p<0.0001; **Table 2**). Significant differences between groups were found for five individual indicators. In the Social Distancing category, maximum occupancy in small enclosed spaces was enforced more in the Lower-prevalence group (Higher-prevalence group 11% vs. Lower-prevalence group 64%, p=0.02; **Table 2**). A greater proportion of LTCFs in the Lower-prevalence group had clear and laminated signage on droplet and contact precaution in required areas (Higher-prevalence group 30% vs. Lower-prevalence group 77%, p=0.03). Notably, a bathroom and sink were present in bedrooms of 100% of LTCFs in the Lower-prevalence group and only in 70% in the Higher-prevalence group (p=0.04). A significantly greater proportion of LTCFs in the Lower-prevalence group conducted trainings and frequent audits to ensure proper mask use among staff members compared to LTCFs in the High-prevalence group (p=0.01). Among the Lower-prevalence group, 100% of LTCFs appropriately used masks inside the COVID-unit and 92% properly used masks outside the COVID-unit. Conversely, appropriate mask use in the Higher-prevalence group was observed in only 50% inside the COVID-unit and 63% outside the COVID-unit. A greater proportion of LTCFs in the Higher-prevalence group had PPE shortages (p=.01) compared to the Lower-prevalence group.

**TABLE 2.** Implementation of infection prevention and control key indicators across Higher- and Lower-prevalence groups.

| ***Key Infection Prevention Indicators*** |  | Total LTCFs<br>Implementing<br>(n=23) | Higher<br>Prevalence<br>Group<br>(n=10) | Lower<br>Prevalence<br>Group<br>(n=13) | P value<sup>a</sup> |
| --- | --- | --- | --- | --- | --- |
| ***Hand Hygiene*** |  |  |  |  |  |
|  | Hand washing training, return demonstrations, and audits frequently conducted with staff? | 18 (78%) | 7 (70%) | 11 (85%) | 0.40 |
|  | Hand sanitizer available at nursing stations, medical carts, in hallways (every 2-3 rooms minimum), and immediately outside of the COVID or observation unit? | 9 (39%) | 2 (20%) | 7 (54%) | 0.11 |
|  | Hand sanitizer available in patient rooms? | 11 (48%) | 5 (50%) | 6 (46%) | 0.85 |
|  | Hand hygiene signage posted throughout facility? | 20 (87%) | 8 (80%) | 12 (92%) | 0.41 |
|  | **Overall Hand Hygiene Implementation** | **63%** | **55%** | **69%** | **0.17** |
| ***Disinfection*** |  |  |  |  |  |
|  | Frequency per day cleaning high-touch areas | mean=4.3 | mean=4.8 | mean=3.9 | 0.46 |
|  | Frequently training staff on cleaning product wet times, auditing of adequate implementation, and implementation of the two-step cleaning method? | 6 (26%) | 2 (20%) | 4 (31%) | 0.56 |
|  | Presence of logs documenting cleaning schedule for shared equipment (IV polls, wheel chairs, IV cuffs, etc.) | 3 (13%) | 2 (20%) | 1 (8%) | 0.41 |
|  | There is a certified IP on staff | 14 (61%) | 5 (50%) | 9 (69%) | 0.37 |
|  | **Overall Cleaning and Disinfection Implementation** | **33%** | **30%** | **36%** | **0.60** |
| ***Social Distancing*** |  |  |  |  |  |
|  | Is there a COVID unit or observation area that is physically separated from COVID-negative residents | 19 (83%) | 8 (80%) | 11 (85%) | 0.76 |
|  | Specific staff assigned to COVID unit or observation area | 18 (78%) | 8 (80%) | 10 (77%) | 0.87 |
|  | Small enclosed areas such as elevators and donning/doffing rooms have signage limiting maximum occupancy | 8/20 (35%) | 1/9 (11%) | 7/11 (64%) | **0.02** |
|  | Droplet and contact precaution signage is laminated and posted outside COVID-unit and individual rooms of COVID-positive residents | 13 (57%) | 3 (30%) | 10 (77%) | **0.03** |
|  | Staff break room is frequently monitored and has adequate space and limited seating to ensure social distancing | 14/22 (64%) | 5 (50%) | 9/12 (75%) | 0.24 |
|  | Resident indoor or outdoor activities, including use of physical therapy or gym facilities have been canceled | 15 (65%) | 6 (60%) | 9 (69%) | 0.66 |
|  | Written out compassionate care policies in place | 11 (48%) | 5 (50%) | 6 (46%) | 0.85 |
|  | Bathroom and sink inside bedroom | 20 (87%) | 7 (70%) | 13 (100%) | **0.04** |

| Overall Social Distancing Implementation |  | 64% | 54% | 72% | 0.01 |
| --- | --- | --- | --- | --- | --- |
| **PPE** |  |  |  |  |  |
|  | Training and frequent audits are conducted to ensure proper mask use by staff members | 14 (61%) | 3 (30%) | 11 (85%) | **0.01** |
|  | Staff members are trained to self-fit test N95 masks | 11 (48%) | 3 (30%) | 8 (62%) | 0.14 |
|  | Masks are stored appropriately if they are being reused | 13/21 (62%) | 5 (50%) | 8/11 (73%) | 0.29 |
|  | Masks are used properly by staff inside the COVID-unit | 18 (78%) | 5 (50%) | 13 (100%) | **<0.01** |
|  | Masks are used properly by staff outside COVID-unit | 18 (78%) | 6 (60%) | 12 (92%) | 0.07 |
|  | Staff are trained and audits take place to ensure proper donning and doffing of PPE | 18 (78%) | 6 (60%) | 12 (92%) | 0.07 |
|  | PPE buddy system is being implemented | 0 (0%) | 0 (0%) | 0 (0%) | 1.00 |
|  | Never had shortage of PPE (past or present) | 13 (57%) | 2 (20%) | 11 (85%) | **<0.01** |
|  | Presence of PPE donning/doffing signage on isolation carts, doors of COVID patients, and at entrance of COVID unit? | 16 (70%) | 7 (70%) | 9 (69%) | 0.96 |
| Overall PPE Implementation |  | 58% | 41% | 72% | **<0.001** |
| Reusing the following |  |  |  |  |  |
|  | Gowns | 9 (39%) | 5 (50%) | 4 (31%) | 0.37 |
|  | N95 Masks | 17 (74%) | 8 (80%) | 9 (69%) | 0.56 |
|  | Surgical Masks | 4 (17%) | 2 (20%) | 2 (15%) | 0.76 |
|  | Face shields | 12 (52%) | 6 (60%) | 6 (46%) | 0.51 |
| Overall PPE Reuse |  | 46% | 53% | 40% | 0.22 |
| **Screening** |  |  |  |  |  |
|  | Frequency of resident temperature screenings per day | mean=3.3 | mean=4.0 | mean=2.85 | 0.10 |
|  | Every entrance to facility includes temperature screening, survey for symptoms and exposures to COVID-19, and 24/7 monitoring of entrance ensuring that hand sanitizer is used | 20 (87%) | 8 (80%) | 12 (92%) | 0.41 |
|  | Temperature and screening logs of staff, residents, and visitors are kept and analyzed daily for trends | 11 (48%) | 3 (30%) | 8 (62%) | 0.14 |
|  | Require 14-day quarantine/observation period for new admissions or readmissions | 21 (91%) | 9 (90%) | 12 (92%) | 0.87 |
| Overall Screening Implementation |  | 75% | 67% | 82% | 0.15 |

<sup>a</sup>Chi square test of proportions used. Significant p values bolded for ease of interpretation.

## Discussion

While studies evaluating IPC implementation in the long-term care setting have been conducted (*8*), there is a dearth of literature that evaluates the implementation of COVID-19 specific IPC recommendations. Through site visits to 23 LTCFs where COVID-19 cases had been reported, we identified IPC categories and specific indicators where implementation was different between Higher- and Lower-prevalence LTCFs. Differences between Higher- and Lower-prevalence LTCFs occurred most frequently in the Social Distancing and PPE categories. Our findings describe the first direct evidence that current recommendations for COVID-19 prevention in LTCFs interrupt transmission and if followed, will reduce the risk of COVID-19 infection among LTCF residents.

One limitation of this analysis is that LTCFs were not selected randomly, but rather based on perceived importance by the FCBOH (prioritizing those with high proportions of resident infection) or per request from a LTCF. These data may not be representative of remaining Fulton County LTCFs that were not identified as “high-priority” or that did not request consultation. A second limitation is that some site visits were done over video-call rather than in person, potentially hindering our ability to observe IPC barriers. Future efforts are being coordinated to conduct in-person follow-up visits to LTCFs visited by video-call, and conduct site visits to remaining Fulton County LTCFs not represented in this analysis.

The COVID-19 pandemic has highlighted the elevated risk of infectious disease outbreaks in LTCFs (*1, 2, 9, 10*). Efforts must be made to build, support, and monitor the capacity of LTCFs to protect the health and safety of residents through strict adherence to IPC recommendations. Guidelines for prevention and control of COVID-19 in LTCFs have been available since the early part of the epidemic (*4-7*). Although they were based on the best available science regarding COVID-19 transmission, there was little data to confirm they indeed protect LTCF residents. Our study provides direct support for these guidelines and suggests that widespread, effective implementation of the guidelines will in fact reduce transmission of COVID-19 in LTCFs.

#### Summary Box

##### What is already known about this topic?

Long-term care facility (LTCF) residents account for a large proportion of infections, hospitalizations, and deaths due to COVID-19.

##### What is added by this report?

Through infection prevention and control (IPC) site visits to 23 LTCFs in Fulton County, Georgia, Higher- and Lower-prevalence groups were compared across IPC categories and indicators. Comparison between the Higher- and Lower-prevalence groups revealed significant differences in PPE and Social Distancing, with five specific indicators driving these differences.

##### What are the implications for public health practice?

Our study provides direct support for the CDC COVID-19 specific IPC guidelines which were provided at the beginning of the COVID-19 pandemic and suggests that widespread, effective implementation will in fact reduce transmission of COVID-19 in LTCFs.

## Data Availability

Data can be accessed through data sharing agreements with the Fulton County Board of Health.

